# Clinico-laboratory profile, intensive care needs, treatment details, and outcome of Pediatric inflammatory multisystem syndrome temporally associated with SARS-CoV-2 (PIMS-TS): A systematic review and Meta-analysis

**DOI:** 10.1101/2020.10.21.20217034

**Authors:** Vijai Williams, Nabaneeta Dash, Renu Suthar, Vichithra Mohandoss, Nishant Jaiswal, TK Kavitha, Karthi Nallasamy, Suresh Kumar Angurana

**Affiliations:** Pediatric Intensive Care Unit, Gleneagles Global Health City, Perumbakkam, Chennai, India.; Pediatric Infectious Diseases Unit, Christian Medical College and Hospital, Vellore, Tamil Nadu, India.; Division of Pediatric Neurology, Department of Pediatrics, Advanced Pediatrics Centre, Postgraduate Institute of Medical Education and Research (PGIMER), Chandigarh, India 160012.; Practising Pediatrician, Chennai, India.; Health Informatics, Department of Telemedicine, Postgraduate Institute of Medical Education and Research (PGIMER), Chandigarh, India 160012.; Division of Pediatric Critical Care, Department of Pediatrics, Advanced Pediatrics Centre, Postgraduate Institute of Medical Education and Research (PGIMER), Chandigarh, India 160012.

**Keywords:** COVID-19, Critically ill children, Hyperinflammation, Intravenous Immunoglobulin, Mechanical Ventilation, MIS-C, Myocarditis, PICU, PIMS-TS, SARS-CoV-2, Steroids

## Abstract

**Objectives:** To synthesize the current data on clinico-laboratory features, intensive care needs, treatment, and outcome of Pediatric inflammatory multisystem syndrome temporally associated with SARS-CoV-2 (PIMS-TS) or multisystem inflammatory syndrome in children (MIS-C).

**Data Sources:** Articles published in PubMed, Web of Science, Scopus, Google Scholar, and WHO COVID-19 research database, CDC database, and Cochrane COVID-19 study register between 1^st^ December 2019 to 10^th^ July 2020.

**Study Selection:** Observational studies involving patients ≤21 years with PIMS-TS or MIS-C, that reported the clinico-laboratory features, intensive care needs, treatment, and outcome.

**Data Extraction:** The search identified 422 citations and finally 18 studies with 833 participants were included and pooled estimate was calculated for parameters of interest utilising random effect model.

**Data Synthesis:** The median age was 9 (8-11) years. Fever, gastrointestinal symptoms, rash, conjunctival injection, and respiratory symptoms were common clinical features. Majority had positive SARS-CoV-2 antibody test and only 1/3^rd^ had RT-PCR positive. The commonest laboratory abnormalities were elevated CRP, D-dimer, procalcitonin, BNP, fibrinogen, ferritin, troponin, and IL-6; and lymphopenia, hypoalbuminemia, and thrombocytopenia. The cardiovascular complications included shock (65%), myocardial dysfunction (61%), myocarditis (65%), and coronary artery abnormalities (39%). Three-fourth children required admission in PICU for mechanical ventilation (25%) and vasoactive drugs (61%). The common treatment provided was IVIG (82%), steroids (54%), antiplatelet drugs (64%), and anticoagulation (51%). The mortality was low (n=13).

**Conclusion:** Fever, gastrointestinal and mucocutaneous symptoms, cardiac dysfunction, shock, and hyperinflammation are common manifestations of PIMS-TS or MIS-C. The short-term outcome is good with supportive intensive care and immunomodulatory treatment.

## Introduction

The coronavirus disease 2019 (COVID-19) pandemic caused by severe acute respiratory syndrome coronavirus 2 (SARS-CoV-2) has affected almost all the countries, overwhelming the healthcare system, and causing significant mortality^.[1-6]^ The severe COVID-19 typically present in 2^nd^ week of illness coinciding with decrease in viral load and increase in inflammatory markers. The host tissue damage is thought to be mediated by dysregulated and aberrant innate and adaptive immune response.^[7-11]^ Acute respiratory failure is the most common organ dysfunction in severe COVID-19 though other organ systems including cardiovascular system may be involved.^[7-9]^ As compared to adults, children are less frequently affected with mild symptoms in majority.^[1-3]^

In mid-April 2020, clinicians from the United Kingdom (UK) reported a cluster of eight previously healthy children who presented with hyperinflammatory shock syndrome which was reported to be temporally associated with COVID-19.^[12]^ The Royal College of Pediatric and Child Health (RCPCH) (1^st^ May 2020), Center for Disease Control and Prevention (CDC) (14^th^ May 2020), and World Health Organization (WHO) (15^th^ May 2020) issued health advisory and criteria to report children presenting with evidence of hyperinflammation and multisystem involvement. Thereafter, multiple reports of pediatric inflammatory multisystem syndrome temporally associated with SARS-CoV-2 (PIMS-TS), multisystem inflammatory syndrome in children (MIS-C), Kawasaki disease (KD), and Kawasaki-like syndrome were published from the countries with high case load of COVID-19 like the UK, France, Italy, and the United States of America (USA) describing the demographic details, clinical features, investigations, treatment details, and outcome.^[12-33]^ The case fatality of COVID-19 in children with PIMS-TS or MIS-C is higher than those without PIMS-TS or MIS-C.

In order to understand the illness and reduce the related morbidity and mortality with PIMS-TS or MIS-C, timely and appropriate information on epidemiology, spectrum of disease, clinical features and course, treatment details, and outcome is needed. This will facilitate development of effective interventions for early diagnosis and treatment as well as scaling up the adequate and effective hospital and intensive care facilities. Therefore, in this systematic review, we described the demographic details, clinical features, laboratory investigations, intensive care needs, management modalities, and outcome of children with PIMS-TS or MIS-C.

## Methodology

This systematic review was conducted as per the Meta-analysis of Observational Studies in Epidemiology (MOOSE) guidelines.^[34]^ The review was registered in PROSPERO (CRD42020198231).

### Search strategy

Three investigators (KN, VW, and SKA) performed independent literature search in electronic databases including PubMed, Web of Science, Scopus, Google Scholar, and WHO COVID-19 research database, CDC database, and Cochrane COVID-19 study register of original articles published between 1^st^ December 2019 and 10^th^ July 2020 using predefined search strategy targeting children and adolescents ≤21 years with PIMS-TS or MIS-C. In addition, preprints from medRxiv and bioRxiv were also screened.

The combination of the following keywords was used as the search strategy for literature search:

- Age group (infants, children, adolescents) with an age restriction of 21 years. AND
- Virus (COVID-19, novel coronavirus, SARS-CoV-2, 2019-nCoV, severe acute respiratory syndrome coronavirus 2) AND
- Condition [PIMS-TS, PIMS, MIS-C, KD, Kawasaki-like syndrome, toxic shock syndrome (TSS), hyperinflammation, hyperinflammatory shock, vasculitis, macrophage activation syndrome (MAS), hemophagocytic lymphohistiocytosis (HLH)].

The references of included studies and review articles were retrieved and screened. Articles published in the English language were included. The Preferred Reporting Items for Systematic Reviews and Meta-analyses (PRISMA) guidelines was followed.^[35]^

### Study Selection

All observational studies involving infants, children, and adolescents upto 21 years, who have been diagnosed with PIMS-TS or MIS-C in association with COVID-19, that reported the demographic profile, clinical features, laboratory investigations, intensive care needs, treatment details, and outcome were considered eligible for this systematic review. Three investigators (VW, KN, and SKA) independently screened the titles and abstracts for the eligibility and later on, all the authors examined the full articles and supplementary contents, if any for inclusion and exclusion criteria.

#### Inclusion criteria

Only studies meeting the following criteria were included:

1. Age group: Infants, children, and adolescents ≤21 years with PIMS-TS or MIS-C in association with COVID-19.
2. Article types: Observational (prospective or retrospective) studies, case series correspondences, brief communications, or letters with data fulfilling data items criteria.
3. Data items: Studies reporting demographical details, clinical features, laboratory investigations, intensive care needs, treatment modalities, and outcome.

#### Exclusion criteria

The following type of studies were excluded:

1. Case reports.
2. Case series reporting <10 cases (The case reports and case series with <10 cases were excluded as these might be part of large studies).
3. Narrative or systematic review.
4. Editorials, letter to editors, correspondences, viewpoints, and opinion letters without original data.
5. Dissertations and conference reports.
6. Other studies that do not meet the inclusion criteria or lack enough data on patient characteristics.

### Data extraction

We used a pre-designed standardized proforma for data extraction. The data entry was be done on Microsoft Excel. Three investigators (VW, KN, and SKA) extracted the data independently from the full text and supplementary contents of the eligible studies. The data collected included first author’s name, journal name and the year of publication, country, study design, number of centres, number of cases, study population, age and sex distribution, method of confirmation of SARS-CoV-2 infection, criteria used to define PIMS-TS or MIS-C, clinical features, laboratory investigations, intensive care needs [PICU admission, mechanical ventilation, vasoactive drugs, renal replacement therapy (RRT), extracorporeal membrane oxygenation (ECMO)], treatment details, and outcome (mortality).

Any disagreement at any point between three investigators was sorted out through discussion and consensus with other two investigators (RS and VM). The data so extracted was rechecked by independent researchers for its accuracy and completeness (ND, KTK, and NJ). To avoid duplicity of the data, efforts were made to screen full text of all included studies for author names, setting, location, date and duration of the study, number of participants, and baseline data.

### Quality assessment

The quality of included studies was assessed using the National Institute of Health Study Quality Assessment Tools for case series studies and observational cohort and cross-sectional studies. The overall risk of bias to each included study was independently assessed by three investigators (VW, KN, and SKA). If any disagreement, other investigators (ND, RS, and VM) were involved to resolve the disagreement. The studies were rated as either having low- or high-risk of bias.

### Main outcome

The outcome of this systematic review was to provide pooled estimate of demographic details, clinical features, laboratory investigations, intensive care needs, treatment details, and outcome (mortality) among children and adolescents (≤21 years) with PIMS-TS or MIS-C.

### Data Synthesis

The initial data entry was done using Microsoft Excel 2013 (Microsoft, Redmond, WA). The descriptive analysis was performed using SPSS version 23 (IBM Corp. 2015. IBM SPSS Statistics for Windows, Version 23.0. Armonk, NY: IBM Corp). The data was presented as number and percentages for categorical variables and median (IQR) for continuous variables.

Meta-analysis was performed by using STATA version 14 (Stata Corp LLC, College Station, Texas, USA). Individual parameters were presented as pooled estimates with a 95% confidence interval (CI) using Metaprop command. The data was pooled from individual studies utilising random effect model with the assumption that the frequency of various parameters was variable across the studies. The statistical heterogeneity among studies was assessed by Chi-Squared test and I^2^ statistics. The heterogeneity was considered to be present if I^2^ >50% and p<0.1.

The subgroup analysis was performed between studies published from the USA and European countries.

## Results

The search identified 422 articles. The 233 duplicate articles and 69 irrelevant articles were removed. Out of 120 full text articles assessed, 102 articles were removed as per the exclusion criteria, and finally 18 articles were included in the final analysis (Figure 1). On quality assessment, eight studies were judged to have had low-risk of bias.^[13, 15-17, 21, 22, 27, 28]^ while 10 had high-risk of bias.^[14, 18-20, 23-26, 29, 30]^ (Table 1)

**Table 1:** Details of studies included in the systematic review

| S. no | Author | Country | No. of centres | No. of cases | Study design | Age group | Criteria used | Study period during 2020 | Risk of bias* |
| --- | --- | --- | --- | --- | --- | --- | --- | --- | --- |
| 1 | Feldstein et al (13) | USA | 53 | 186 | Retrospective and Prospective | 0-21 years | CDC | 15-Mar to 20-May | Low |
| 2 | Belot et al (14) | France | 7 | 108 | Retrospective and Prospective | 0-15 years | RCPCH | 01-Mar to 17-May | High |
| 3 | Dufort et al (15) | USA | 106 | 99 | Retrospective | 0-21 years | NYSDOH | 01-Mar to 10-May | Low |
| 4 | Davies et al (16) | UK | 21 | 78 | Retrospective and prospective | 0-18 years | RCPCH | 01-Apr to 10-May | Low |
| 5 | Whittaker et al (17) | UK | 8 | 58 | Retrospective | 0-18 years | RCPCH | 23-Mar to 16-May | Low |
| 6 | Miller et al (18) | USA | 1 | 44 | Retrospective | 0-21 years | CDC | 18-Apr to 22-May | High |
| 7 | Hameed et al (19) | UK | 1 | 35 | Retrospective | 4-14 years | RCPCH | 14-Apr to 09-May | High |
| 8 | Belhadjer et al (20) | France and Switzerland | 13 | 35 | Retrospective | 2-16 years | Fever, cardiogenic shock, or acute left ventricular dysfunction with inflammatory state | 22-Mar to 30-Apr | High |
| 9 | Capone et al (21) | USA | 1 | 33 | Retrospective | 0-21 years | CDC | 17-Apr to 13-May | Low |
| 10 | Kaushik et al (22) | USA | 3 | 33 | Retrospective | 0-21 years | CDC | 03-Apr to 23-May | Low |
| 11 | Toubiana et al (23) | France | 1 | 21 | Prospective | 0-18 years | AHA KD | 27-Apr to 11-May | High |
| 12 | Grimaud et al (24) | France | 4 | 20 | Retrospective | 0-18 years | Fever, shock, acute myocarditis | 15-Apr to 27-Apr | High |
| 13 | Cheung et al (25) | USA | 1 | 17 | Retrospective | 0-21 years | AHA KD | 18-Apr to 05-May | High |
| 14 | Pouletty et al (26) | France | 7 | 16 | Retrospective | 0-18 years | AHA KD | 1 April to 30 April | High |
| 15 | Riollano-Cruz et al (27) | USA | 1 | 15 | Retrospective | 0-21 years | CDC and NYSDOH | 24-Apr to 14-May | Low |
| 16 | Ramcharan et al (28) | UK | 1 | 15 | Retrospective |  | RCPCH | 10-Apr to 09-May | Low |
| 17 | Ouldeli et al (29) | France | 1 | 10 | Retrospective | 1.5-15.8 years | AHA KD | 04-May to 28-Apr | High |
| 18 | Verdoni et al (30) | Italy | 1 | 10 | Retrospective | 3-16 years | AHA KD | 18-Feb to 20-Apr | High |

\*Study quality was assessed using National Institute of Health Study Quality Assessment Tools for case series studies and observational cohort and cross-sectional studies.
AHA: American Heart Association, CDC: Centers for Disease Control and Prevention, KD: Kawasaki Disease, NYSDOH: New York State Department of Health, RCPCH: Royal College of Pediatric and Child Health, UK: United Kingdom, US: United States of America.

**Figure 1:**
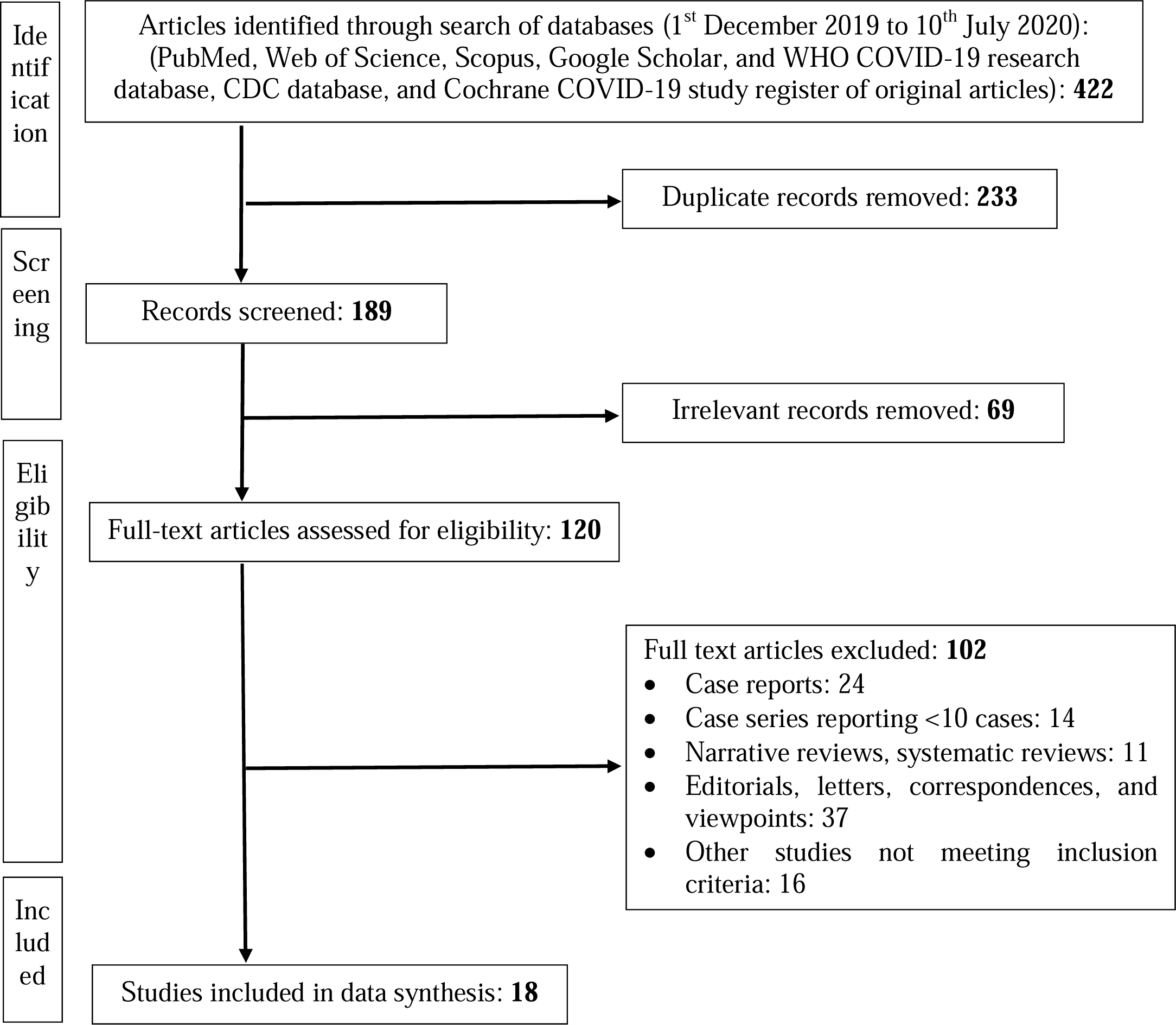
Flow diagram of study selection

### Study characteristics

In 18 studies, 833 children were reported. All the studies were conducted between 1^st^ March and 23^rd^ May 2020 with median (IQR) length of the study duration being 39 (24-56) days. Seven studies were from the USA^[13, 15-18, 21, 22, 25]^ and 11 were from Europe^[14, 16-17, 19, 20, 23, 26, 28-30]^ with almost 50% cases from each region. The studies from Europe include 4 studies from UK^[16, 17, 19, 28]^, 5 from France^[14, 23-24, 26, 29]^, 1 from France and Switzerland^[20]^, and 1 from Italy.^[30]^ Nine studies were single centre^[18, 19-21, 23, 25,-30]^ and 9 were multicentre studies.^[13-17, 20-22, 24, 26]^ The study design was retrospective in 14 studies,^[15, 17-22, 24-30]^ retrospective and prospective in 3,^[13, 14, 16]^ and prospective in 1 study.^[23]^ The criteria used to define the inflammatory syndrome in children in temporal relation to COVID-19 was CDC and/or New York State Department of Health (NYSDOH) criteria in 6 studies,^[13, 15-18, 21, 22,]^ RCPCH criteria in 5 studies,^[14, 16-17, 19, 28]^ and American Heart Association (AHA) KD criteria in 5 studies.^[23, 25-26, 29, 30]^ Two studies enrolled cases with fever, shock, acute myocarditis/left ventricular (LV) dysfunction, and evidence of inflammation.^[20, 24]^ (Table 2)

**Table 2:** Characteristics of the included studies.

| **Study characteristics** | **Studies, n (%)** | **Cases, n (%)** |
| --- | --- | --- |
| **Included in review** | 18 (100) | 833 (100) |
| **Region** |  |  |
| USA | 7 (38.9) | 427 (51.3) |
| Europe | 11 (61.1) | 406 (48.7) |
| **Country** |  |  |
| USA | 7 (38.9) | 427 (51.3) |
| UK | 4 (22.2) | 186 (22.3) |
| France | 5 (27.8) | 175 (21) |
| France and Switzerland | 1 (5.6) | 35 (4.2) |
| Italy | 1 (5.6) | 10 (1.2) |
| **Number of centres** |  |  |
| Single centre | 9 (50) | 200 (24) |
| Multiple centres | 9 (50) | 633 (76) |
| **Study design** |  |  |
| Retrospective | 14 (77.8) | 440 (52.8) |
| Retrospective and prospective | 3 (16.7) | 372 (44.7) |
| Prospective | 1 (5.6) | 21 (2.5) |
| Study duration in days, median (IQR) | 39 (24-56) |  |
| **Criteria used** |  |  |
| CDC and/or NYSDOH | 6 (33.3) | 410 (49.2) |
| RCPCH | 5 (27.8) | 294 (35.5) |
| Kawasaki disease (AHA) | 5 (27.8) | 74 (8.9) |
| Fever, shock, acute myocarditis/LV dysfunction, and inflammation | 2 (11.1) | 55 (6.6) |

There was overlap of few cases in studies from UK,^[16, 17, 19, 28]^ France,^[26, 29]^ and USA.^[22, 27]^ As the information to identify overlapping cases could not be derived, after discussion among 3 authors (VW, KN, and SKA), consensus was reached to include all these studies in the systematic review.

### Clinical features

All except one study (15) reported median (IQR) age and it was 9 (8-11) years. All studies reported gender and males were 57%. The median (IQR) duration of illness/fever was 5 (4-6) days as reported in 11 studies.^[13, 16, 20-26, 28, 30]^ Race was reported in 12 studies^[13, 15-18, 21-23, 25, 26, 28, 30]^ and common races noted were black (35%), white (27%), Asian (10%), and others (14%). All except 4 studies^[14, 19, 28, 29]^ reported comorbidities (29%); common comorbidities noted were underlying respiratory, cardiac, immunocompromising, autoimmune diseases, and obesity (Table 3).

**Table 3:** Clinical features in children with inflammatory syndrome in association with SARS-CoV-2 infection.

| **Characteristics** | **Number of studies** | **Number of cases** | **Pooled estimate % (95% CI)** | **Heterogeneity ($I^2$ %)** |
| --- | --- | --- | --- | --- |
| Age in years, Median (IQR) | 17 | 734 | 9 (8-11) |  |
| Duration of illness in days, Median (IQR) | 11 | 725 | 5 (4-6) |  |
| Male gender | 18 | 833 | 57 (51-62) | 88* |
| **Race** | 12 | 610 |  |  |
| Black |  |  | 35 (28-42) | 69* |
| White |  |  | 27 (10-35) | 83* |
| Other |  |  | 14 (6-22) | 84* |
| Asian |  |  | 10 (5-15) | 76* |
| Comorbidity | 14 | 665 | 29 (22-36) | 74* |
| **Clinical features** |  |  |  |  |
| Fever | 17 | 725 | 96 (89-100) | 89* |
| Gastrointestinal symptoms | 16 | 715 | 86 (82-90) | 51* |
| Rash | 14 | 667 | 58 (52-65) | 63* |
| Conjunctival injection | 13 | 632 | 52 (40-63) | 89* |
| Respiratory symptoms | 8 | 445 | 43 (27-59) | 91* |
| Oral mucosal changes | 10 | 504 | 42 (29-55) | 78* |
| Peripheral extremity changes | 6 | 306 | 39 (24-52) | 82* |
| Neurological symptoms | 11 | 529 | 32 (10-42) | 82* |
| Cervical adenopathy | 9 | 462 | 24 (14-34) | 88* |
| Musculoskeletal symptoms | 5 | 337 | 17 (10-24) | 55* |
| **Confirmation of exposure** |  |  |  |  |
| SARS-CoV-2 RT-PCR and/or antibody positive | 14 | 722 | 85 (74-91) | 89* |
| SARS-CoV-2 antibody test positive | 18 | 833 | 84 (72-93) | 89* |
| SARS-CoV-2 RT-PCR positive | 18 | 833 | 37 (28-46) | 89* |

\*P value for $I^2 < 0.1$

Fever was the most common symptom reported in 96% children in all except one study.^[14]^ Gastrointestinal symptoms (pain abdomen, nausea/vomiting, and diarrhea) were noted in 86% in all expect 2 studies.^[14, 29]^ Other common clinical features noted were rash (58%), conjunctival injection (52%), respiratory symptoms (43%), oral mucosal changes (42%), peripheral extremity changes (39%), neurological symptoms (32%), cervical lymphadenopathy (24%), and musculoskeletal symptoms (17%) (Table 3).

The results of SARS-CoV-2 antibody test and RT-PCR were reported by all studies and these were positive in 84% and 37% children, respectively (Table 3).

### Laboratory investigations

The laboratory investigations are shown in Supplemental table 1. Radiological investigations were reported in 11 studies^[13, 15-18, 19, 22, 23-28, 30]^ and the findings are described in Supplemental table 2. Lymphopenia was noted in 85% and thrombocytopenia in 53% children. Among inflammatory markers, elevated CRP was noted in 98% children, elevated procalcitonin in 90%, elevated fibrinogen in 86%, elevated ferritin in 82%, and elevated IL-6 in 68% children. Elevation of D-dimer was noted in 92% children, hypoalbuminemia in 71%, and elevated ALT in 40%. Among cardiac markers, 89% children had elevated BNP (NT-BNP or Pro-BNP) and 78% had elevated troponin (Table 4).

**Table 4:** Laboratory abnormalities, echocardiographic findings and other cardiovascular manifestations.

| **Characteristics** | **Number of studies** | **Number of cases** | **Pooled estimate % (95% CI)** | **Heterogeneity ($I^2$ %)** |
| --- | --- | --- | --- | --- |
| Lymphopenia | 10 | 514 | 85 (76-93) | 50* |
| Thrombocytopenia | 4 | 330 | 53 (43-98) | 96* |
| Elevated CRP | 13 | 603 | 98 (79-100) | 93* |
| Elevated procalcitonin | 7 | 221 | 90 (76-100) | 82* |
| Elevated fibrinogen | 5 | 330 | 86 (73-96) | 85* |
| Elevated ferritin | 8 | 477 | 82 (67-98) | 88* |
| Elevated IL-6 | 6 | 178 | 68 (32-98) | 74* |
| Elevated D-dimer | 8 | 477 | 92 (82-100) | 97* |
| Hypoalbuminemia | 7 | 401 | 71 (50-93) | 91* |
| Elevated ALT | 7 | 330 | 40 (23-56) | 86* |
| Elevated BNP (any) | 11 | 510 | 89 (82-96) | 90* |
| Elevated troponin | 12 | 371 | 78 (63-93) | 86* |
| Thrombosis | 3 | 297 | 2 (2-7) | 25* |
| Arrhythmia | 7 | 347 | 7.2 (2.5-13.3) | 30 |
| **Echocardiography performed** | 16 | 681 | 94 (91-96) | 44 |
| Myocardial dysfunction or ejection fraction <55% | 15 | 603 | 61 (50-72) | 77* |
| Myocarditis (clinical and/or biochemical and/or echocardiography) | 12 | 588 | 65 (49-80) | 87* |
| Any coronary artery abnormality | 9 | 248 | 39 (25-63) | 90* |
| Coronary artery dilatation or aneurysm | 16 | 681 | 16 (12-20) | 51* |
| Coronary artery diameter >+2.5 z score | 12 | 551 | 8 (6-11) | 16 |
| Pericardial effusion | 12 | 477 | 35 (24-46) | 74* |
| Residual myocardial dysfunction at discharge | 8 | 179 | 10.4 (2.5-18.3) | 20 |

\*P value for $I^2 < 0.1$

### Echocardiography findings

Sixteen studies reported data on echocardiography which was done in 94% children.^[13, 15-18, 20-30]^ The common echocardiographic abnormalities include LV dysfunction or ejection fraction <55% (61%), myocarditis (as defined by clinical and/or biochemical and/or echocardiographic diagnosis) (65%), any coronary artery abnormality (39%), pericardial effusion (35%), coronary artery dilatation or aneurysm (16%), and coronary artery diameter >2.5 z-score (8%) children. About 10% children had residual LV dysfunction at discharge as reported in eight studies.^[20-22, 24-26, 28, 30]^ (Table 4)

### Intensive care needs

All studies reported intensive care admission and 76% children were managed in PICU where they received high flow nasal cannula oxygen (18%), non-invasive ventilation (22%), and invasive ventilation (25%). Shock was noted in 65% children and 61% required vasoactive/inotropic drugs. Acute kidney injury was noted in 35% children and 2% underwent RRT. The requirement of ECMO was reported in all except 3 studies^[14, 26, 30]^ and it was used in 4% children (n=32) (Table 5).

**Table 5:** Intensive care needs, treatment details, and outcome.

| Characteristics | Number of studies | Number of cases | Pooled estimate % (95% CI) | Heterogeneity ($I^2$ %) |
| --- | --- | --- | --- | --- |
| Admission in PICU | 18 | 833 | 76 (65-88) | 80* |
| HFNC | 5 | 389 | 18 (7-29) | 73* |
| Non-invasive ventilation | 9 | 497 | 22 (13-31) | 84* |
| Invasive ventilation | 18 | 833 | 25 (19-37) | 92* |
| Shock | 18 | 833 | 65 (54-73) | 88* |
| Vasoactive drugs | 18 | 833 | 61 (53-70) | 86* |
| Acute kidney injury | 8 | 477 | 35 (21-50) | 95* |
| Renal replacement therapy | 5 | 363 | 2 (1-4) | 65* |
| ECMO | 15 | 699 | 4 (1-8) | 33 |
| IVIG | 16 | 690 | 82 (74-89) | 65* |
| 2<sup>nd</sup> dose IVIG (among those who received 1<sup>st</sup> dose) | 8 | 326 | 25 (11-33) | 83* |
| Steroids | 16 | 690 | 54 (41-67) | 94* |
| Steroids + IVIG | 4 | 306 | 50 (39-62) | 67* |
| IL-6 inhibitors (Tocilizumab or siltuximab) | 13 | 533 | 12.6 (1-26) | 88* |
| IL-1Ra inhibitor (Anakinra) | 12 | 549 | 8.6 (5-12) | 0.00 |
| Infliximab | 9 | 469 | 2.9 (0.1-6.8) | 54* |
| Rituximab | 6 | 210 | 0.2 (0-0.8) | 43 |
| Plasma therapy | 7 | 333 | 2 (0-4) | 24 |
| Anticoagulation | 11 | 487 | 51 (25-77) | 93* |
| Antiplatelets (any dose) | 9 | 238 | 64 (30-78) | 95* |
| Discharged at the time of reporting | 17 | 725 | 95 (91-100) | 92* |
| Still hospitalized at the time of reporting | 17 | 725 | 3.6 (0.5-7.8) | 88* |
| Deaths at the time of reporting | 18 | 833 | 2 (1-3) | 44 |
| Duration of stay, median (IQR) | 11 | 376 | 7.8 (5-10) |  |

\*P value for $I^2 < 0.1$

### Treatment details

All except two studies^[14, 19]^ reported data on intravenous immunoglobulin (IVIG) and steroids use that were given to 82% and 54% children, respectively. The second dose of IVIG was used in 25% of children who did not show improvement after the first dose.^[13, 20-21, 23, 26,-30]^ The combination of steroid plus IVIG was used in 50% children.^[13, 15, 23, 30]^ Other biological and immunomodulator agents used were IL-6 inhibitors (12.6%) and IL-1Ra inhibitor (8.6%). Sixteen children received infliximab, 1 rituximab, and 5 received convalescent plasma therapy. Anticoagulation (prophylactic or therapeutic) and antiplatelet agents (any dose) were used in 51% and in 64% children, respectively (Table 5).

### Outcome

All studies reported the number of deaths. There were 13 deaths with pooled estimate% (95% CI) of 2 (1-3). The data on hospital discharge was given in all except one study.^[14]^ Majority of children (95%) were discharged at the time of reporting and only 3.6% were still hospitalized (n=77). The median (IQR) duration of hospital stay was 7.8 (5-10) days (Table 5).

### Sub-group analysis

We compared the studies from the USA and Europe region. Significant differences between two regions include differences in criteria used (USA: CDC and/or NYSDOH criteria; Europe: RCPCH and AHA KD criteria); higher proportion of children with comorbidity (36% vs. 19%, p=0.04), and greater frequency of treatment with IL-6 inhibitors (27% vs. 4%, p=0.02) and IL-1Ra inhibitors (13% vs. 5%, p=0.004) in studies from the USA; and higher need of mechanical ventilation in studies from Europe (33% vs. 12%, p=0.03) (Supplemental table 3).

## Discussion

In this systematic review, we summarized the epidemiological and clinical features, investigations, intensive care needs, treatment details, and outcome of children and adolescents (<21 years) with PIMS-TS or MIS-C. This illness is characterized by febrile hyperinflammatory state with gastrointestinal, mucocutaneous, dermatological, and cardiac manifestations. The peak incidence occurred about a month after the peak of COVID-19 epidemic (when COVID-19 activity was decreasing) and then the incidence decreased thereafter.^[13-16, 30]^ As several countries are progressing toward the peak of COVID-19 pandemic, the number of children presenting with PIMS-TS or MIS-C would increase with a possible surge in countries where this syndrome has not been reported till now. These countries should prepare themselves to manage the surge in children with this syndrome with in next few weeks after the peak of the COVID-19 pandemic. In this context, this systematic review describing clinico-epidemiological features, management options, and outcome of children with PIMS-TS or MIS-C assumes significance in sensitizing about this delayed but life-threatening complications of SARS-CoV-2 infection which otherwise causes mild illness in majority of children and adolescents.

The possible pathogenesis suggested for the development of PIMS-TS or MIS-C are immune-mediated injury and inflammatory vasculopathy triggered by SARS-CoV-2 infection rather than active viral infection.^[11, 13, 15]^ The facts supporting this hypothesis are onset of symptoms 2-4 weeks after SARS-CoV-2 infection; majority of children had laboratory evidence of recent or concurrent SARS-CoV-2 infection in form of positive SARS-CoV-2 antibody test or SARS-CoV-2 RT-PCR suggesting a temporal association between SARS-CoV-2 and PIMS-TS or MIS-C; an exuberant host inflammatory response; and potential benefit with immunomodulatory drugs (IVIG and/or steroids).

We noted that most children were older (median age 9 years), previously healthy, and presented 4-6 weeks after the peak of COVID-19. The proportion of children belonged to black race were higher (35%) similar to what was noted in adults with severe clinical presentations of COVID-19 for which a possible genetic predisposition needs to be explored.^[36]^ The most common presenting symptoms were fever, GI symptoms, rash, conjunctival injection, respiratory symptoms, and oral mucosal changes. GI symptoms (86%) were strikingly prominent.^[13, 15-22, 24-26, 28, 30]^ mimicking GI infection, acute abdomen, or inflammatory bowel disease.^[18, 19, 37]^ The possible mechanisms for the GI symptoms could be bowel wall edema or ischemia due to vasculitis, cardiac dysfunction and/or shock, mesenteric inflammation, and mesenteric lymphadenitis.^[19, 23, 37]^

The symptom complex of fever, GI symptoms, and rash in children with SARS-CoV-2 infection (symptomatic or asymptomatic) in recent past (2-4 weeks prior) should alert clinicians to early recognize this syndrome, prompt investigation for hyperinflammation and organ dysfunction, close monitoring (including hemodynamic monitoring, electrocardiography, and echocardiography), and aggressive supportive and specific therapy.

Majority of children had elevated inflammatory markers (CRP, procalcitonin, fibrinogen, ferritin, D-dimer, IL-6), lymphopenia, hypoalbuminemia, and thrombocytopenia. These hyperinflammatory manifestations noted were similar to adults with COVID-19.^[38, 39]^

Among cardiac manifestations, 65% children had evidence of myocarditis, 61% had evidence of LV dysfunction, 16% had coronary dilatation or aneurysm with 8% documented coronary artery diameter >2.5 z-score. About 10% children had residual LV dysfunction at discharge. In adults with COVID-19, myocardial dysfunction has been observed to be a prominent extrapulmonary manifestation that was associated with increased mortality.^[40, 41]^

The common chest radiological abnormalities noted in children with COVID-19 are bronchial thickening, ground-glass opacities, or inflammatory lung lesions, suggestive of pneumonia. These lung findings were also noted in asymptomatic children and those with mild symptoms, suggesting that SARS-CoV-2 infection induces a primary inflammation of lung parenchyma and lower respiratory tract.^[42, 43]^ The reporting of radiological features was not uniform in included studies.

Most children with acute SARS-CoV-2 infections are usually asymptomatic or having mild symptoms,^[1-4]^ while majority with PIMS-TS or MIS-C were noted to have severe disease requiring PICU admission (76%), vasoactive drugs (61%), and invasive mechanical ventilation (25%). The most common treatment strategies used were IVIG (82%) and steroids (54%). Small proportion of children also received IL-6 inhibitors, IL-1Ra inhibitor, infliximab, rituximab, and convalescent plasma therapy. Antiplatelets and anticoagulation were used in 64% and 51% children, respectively. The short-term morbidity was high in terms of higher requirement of intensive care interventions, but the mortality was low.

The presentation of PIMS-TS or MIS-C had some overlapping features with KD, TSS, HLH, or MAS.^[15, 16]^ However, it differed from KD on following accounts: older age at presentation (older children and adolescents); higher proportion of children with GI and respiratory symptoms; predominance of severe cardiovascular system involvement in form of shock, LV dysfunction, or myocarditis; higher proportion of lymphopenia, thrombocytopenia, and elevated CRP and procalcitonin; and more children being cared in PICU and requiring vasoactive drugs and mechanical ventilation.^[11-13, 15-16, 20, 30]^

As per the available data, it seems that PIMS-TS or MIS-C is an uncommon manifestation of SARS-CoV-2 infection reported at greater frequency among specific age group, ethnicity (blacks), and region (scarcity of reports from Asia including China). These differences could be due to differences in exposure to SARS-CoV-2 infection, incomplete reporting, non-severe disease, predominance of SARS-CoV-2 infection among black race, differences in nasal expression of angiotensin-converting enzyme 2 (ACE2) receptors for SARS-CoV-2 cell entry; socioeconomic status, and comorbidities. The susceptibility to inflammatory disease and response to treatment may also be influenced by the gut microbiome, signalling pathways, genetic variations and other host factors; and early treatment with immunomodulators.^[9, 13-16, 44, 45]^

There is great variation among clinicians in the use of immunomodulatory treatments of PIMS-TS or MIS-C and IVIG and steroids were used most commonly. However, good quality evidence from well-designed clinical trials are required to establish treatment guidelines. In absence of definite evidence of specific treatment, the supportive intensive care and multidisciplinary approach (intensivist, infectious disease specialist, cardiologist, haematologist, immunologist/rheumatologist, and pharmacologist) remains crucial for clinical management.

This systematic review has several strengths. To the best of our knowledge, this is the first review that summarized the available literature on epidemiology, clinical features, investigations, intensive care needs, treatment, and outcome of children with PIMS-TS or MIS-C. The search strategy was rigorous to include all studies that reported children with hyperinflammatory syndrome associated with COVID-19 irrespective of the description (PIMS-TS, MIS-C, KD, cardiac involvement, acute heart failure, acute myocarditis, GI manifestations, or imaging findings). We also performed a sub-group analysis to compare studies from two continents (USA and Europe) which did not demonstrate significant differences in characteristics despite ethnic differences.

The systematic review also has several limitations. All the included studies were conducted over a short period of time. Most of the studies were retrospective and with small sample size. More than half of the studies had high-risk of bias. The criteria used were different across the region. There was a small overlap of few cases in few studies which we could not delineate. The long-term follow-up data was not available which is needed to identify long-term health issues (especially those with myocardial dysfunction and coronary artery abnormalities). We do not have studies from other countries with high burden of COVID-19 (Brazil, India, Russia, South Africa, Mexico, Spain etc.) at the time this review was performed which could possibly contribute to publication bias.

## Conclusion

The children with PIMS-TS or MIS-C were reported from the USA and Europe regions 4-6 weeks after the peak of COVID-19 pandemic. Most of the affected children were previously healthy and had laboratory evidence of recent or concurrent SARS-CoV-2 infection. Fever, GI symptoms, rash, and mucocutaneous manifestations were commonest clinical features. Majority of children had evidence of systemic inflammation, cardiovascular involvement, required PICU admission, and treated with vasoactive drugs and immunomodulators (IVIG and/or steroids). The short-term outcome was good with low mortality.

## Data Availability

Data will be available

