## Supplemental Tables 1-3 for "Clinico-laboratory profile, intensive care needs, treatment details, and outcome of Pediatric inflammatory multisystem syndrome temporally associated with SARS-CoV-2 (PIMS-TS): A systematic review and Meta-analysis"

**Supplemental Table 1: Laboratory investigations.**

| **Characteristics** | **Number of studies** | **Number of cases** | **Median (IQR)** |
| --- | --- | --- | --- |
| Hemoglobin (g/dL), median (IQR) | 7 | 216 | 11 (9-11) |
| Platelet count (×10⁹ cells/L), median (IQR) | 14 | 660 | 168 (146-216) |
| Leucocyte count (×10⁹ cells/L), median (IQR) | 10 | 343 | 11 (10-15) |
| Lymphocyte count (×10⁹ cells/L), median (IQR) | 12 | 514 | 1.04 (0.8-1.1) |
| ESR (mm/hr) | 6 | 387 | 63 (57-72) |
| CRP (mg/L) | 15 | 693 | 229 (147-251) |
| Procalcitonin (ng/ml) | 8 | 274 | 17 (8-33) |
| Ferritin (ng/ml) | 11 | 580 | 639 (560-1042) |
| Fibrinogen (mg/dL) | 6 | 253 | 625 (608-724) |
| D-dimer (ng/ml) | 12 | 620 | 3899 (2695-4075) |
| Albumen (g/dL) | 10 | 520 | 2.8 (2.1-3.4) |
| ALT (IU/L) | 10 | 324 | 40 (33-55) |
| LDH (U/L) | 4 | 234 | 324 (314-396) |
| IL-6 (pg/ml) | 9 | 290 | 200 (153-240) |
| BNP (pg/ml) | 5 | 295 | 3354 (791-4574) |
| NT-BNP (pg/ml) | 8 | 217 | 4322 (1773-22323) |
| Troponin (ng/ml) | 12 | 371 | 96 (50-279) |

**Supplemental Table 2: Description of radiological investigations.**

| **S. no.** | **Author** | **Number of cases** | **Imaging findings** |
| --- | --- | --- | --- |
| 1 | Feldstein et al (13) | 186 | Chest radiograph revealed infiltrates in 42% (n=72) and pleural effusion in 27% (n=50) children.  Abdominal ultrasonography showed gall bladder hydrops in 2% (n=3) children. |
| 2 | Dufort et al (15) | 99 | Chest computed tomography (CT) or chest radiograph was done in 91% (n=90) children and opacities noted in 39% (n=35) children.  Abdomen CT, ultrasonography, or both were done in 44.4% (n=44) children and these were abnormal in 77% (n=34). The common abnormalities were hepatomegaly, splenomegaly, or hepatosplenomegaly (9%), mesenteric adenopathy (18%), ascites, pleural effusions, or pelvic fluid (36%), and inflammation or enlargement of appendix (4.5%), gallbladder (11.4%), enteritis or enterocolitis (6.8%), bowel-wall thickening (15.9%), or fluid filled bowel loops (9%). |
| 3 | Miller et al (18) | 44 | Abdominal imaging was performed in 34.1% (15/44) children and these were abnormal in 80% (n=12) children.  The common findings were mesenteric adenitis (n=2), biliary sludge or acalculous cholecystitis (n=6), and ascites (n=6). |
| 4 | Hameed et al (19) | 35 | Chest radiographs were abnormal in 54% (n=19) children and common abnormalities were peribronchial cuffing and perihilar interstitial thickening, consolidation, pleural effusion, and atelectasis.  CT chest was performed in 94% (n=33) children and common findings noted were basal consolidation with collapse (39%, n=13), pleural effusions (30%, n=10), and diffuse bilateral ground-glass (9%, n=3).  The abdominal ultrasonography was performed in 54% (n=19) children and it was abnormal in 95% (n=18) children. The common findings were anechoic free-fluid (53%), localised inflammatory change within the right iliac fossa (47%), echogenic expanded mesenteric fat (37%), multiple mildly enlarged lymph nodes (47%), bowel wall thickening (21%), increased periportal echogenicity (16%), pericholecystic oedema and mild gallbladder wall thickening (16%), and gallbladder sludge (16%).  The common CT abdomen findings (n=5) noted were mesenteric fat-stranding and lymphadenopathy in right iliac fossa (60%, n=3), marked distal ileal and cecal bowel wall thickening (n=1), free fluid (n=4), and splenic infarct (n=1). |
| 5 | Kaushik et al (22) | 33 | Chest radiographs revealed cardiomegaly in 30% (n=10), focal opacity in 15% (n=5), and bilateral opacities in 18% (n=6) children. |
| 6 | Toubiana et al (23) | 21 | Chest imaging (radiography or computed tomography) was abnormal in 44% (8/18) children and common abnormalities noted were ground glass opacities, local patchy shadowing, and interstitial abnormalities. |
| 7 | Cheung et al (25) | 17 | Abnormal chest radiograph was noted in 82.3% (14/17) children.  One child had acute ileocolitis on abdominal imaging. |
| 8 | Pouletty et al (26) | 16 | Chest radiograph was abnormal in 31% (n=5) children. |
| 9 | Riollano-Cruz et al (27) | 15 | Chest radiographs revealed lung opacities or ground glass opacities in 47% (n=7), reactive airway disease in 27% (n=4), and pleural effusion in 27% (n=4) children. |
| 10 | Ramcharan et al (28) | 15 | Chest radiographs were performed in 93.3% (n=14) children and abnormal in 50% (n=7). The common abnormalities were pleural effusions (35.7%), consolidation (21.4%), and cardiomegaly (14.3).  The abdominal ultrasonography in 6 children did not revealed any abnormality. |
| 11 | Verdoni et al (30) | 10 | Chest radiographs showed infiltrates in 50% (n=5) children.  Two children underwent CT chest and noted to have bibasilar pulmonary thickening. |

**Supplemental Table 3: Characteristics of the included studies and children from USA and Europe regions.**

| **Parameter** | **USA*** | **Europe*** | **p** |
| --- | --- | --- | --- |
| **Study characteristics** |  |  |  |
| Number of studies, n (%) | 7 (38.9) | 11 (61.1) |  |
| Total cases, n (%) | 427 (51.3) | 406 (48.7) | 0.76 |
| Study duration (days), median (IQR) | 34 (20-66) | 35 (25-54) | 0.93 |
| Number of centres, n (%) |  |  |  |
| Single centre, n (%)  Multiple centres, n (%) | 4 (57.1)  3 (42.9) | 5 (45.5)  6 (54.5) | 0.5 |
| **Criteria used** |  |  |  |
| CDC and/or NYSDOH, n (%)  RCPCH, n (%)  Kawasaki disease (AHA), n (%)  Fever, shock, acute myocarditis or  LV dysfunction, and inflammation, n (%) | 6 (85.7)  0  1 (14.3)  0 | 0  5 (45.5)  4 (36.4)  2 (18.2) | **0.002** |
| **Population characteristics** |  |  |  |
| Age in years, Median (IQR) | 8.5 (8-11) | 10 (8-27) | 0.54 |
| Male gender | 58 (49-67) | 56 (47-65) | 0.65 |
| Comorbidity | 36 (27-45) | 19 (10-29) | **0.04** |
| Fever | 94 (90-100) | 98 (92-100) | 0.79 |
| Gastrointestinal symptoms | 87 (81-93) | 86 (82-90) | 0.75 |
| Rash | 59 (52-66) | 58 (47-69) | 1 |
| Respiratory symptoms | 44 (27-62) | 43 (27-59) | 0.73 |
| **Confirmation of exposure** |  |  |  |
| RT PCR and/or antibody positive | 89 (86-93) | 82 (73-91) | 0.12 |
| Antibody test positive | 86 (68-100) | 82 (70-94) | 0.29 |
| RT PCR positive | 43 (35-50) | 34 (20-45) | 0.17 |
| **Investigations** |  |  |  |
| Lymphopenia | 75 (65-85) | 84 (67-93) | 0.09 |
| Thrombocytopenia | 48 (30-67) | 58 (11-80) | 1.0 |
| Elevated CRP | 98 (91-99) | 98 (79-100) | 0.82 |
| Elevated procalcitonin | 83 (72-91) | 100 (88-100) | 0.16 |
| Elevated ferritin | 74 (48-95) | 87 (68-100) | 0.29 |
| Elevated D-dimer | 90 (65-100) | 94 (79-100) | 0.76 |
| Elevated BNP (any) | 88 (70-100) | 90 (80-100) | 0.63 |
| Elevated troponin | 68 (27-98) | 83 (61-100) | 0.36 |
| **Echocardiographic findings** |  |  |  |
| Myocardial dysfunction or EF <55% | 58 (43-64) | 63 (37-80) | 0.91 |
| Myocarditis | 48 (13-81) | 70 (52-90) | 0.31 |
| Any coronary artery abnormality | 42 (27-56) | 37 (9-66) | 0.33 |
| Coronary artery dilatation or aneurysm | 12 (7-17) | 19 (14-23) | 0.79 |
| Coronary artery diameter >2.5 z score | 7 (4-11) | 12 (7-18) | 0.94 |
| Pericardial effusion | 38 (27-48) | 21 (13-41) | 0.22 |
| Residual myocardial dysfunction at discharge | 13 (4-21) | 9 (5-16) | 0.65 |
| **Intensive care needs** |  |  |  |
| Admission in PICU | 88 (78-97) | 69 (52-86) | 0.07 |
| Invasive ventilation | 12 (6-21) | 33 (24-46) | **0.03** |
| Shock | 61 (47-75) | 66 (56-76) | 0.65 |
| Vasoactive drugs | 57 (48-66) | 62 (47-77) | 0.59 |
| AKI | 25 (7-40) | 50 (30-66) | 0.19 |
| RRT | 2.5 (1.5-8.8) | 0.3 (0.1-1.2) | **0.07** |
| ECMO | 2.6 (0-4.3) | 5.4 (2-6) | 0.86 |
| **Treatment details** |  |  |  |
| IVIG | 77 (70-82) | 85 (70-91) | 0.42 |
| 2^nd^ dose IVIG (among those who received 1^st^ dose) | 30 (20-44) | 23 (5-36) | 0.40 |
| Steroids | 63 (44-82) | 46 (29-64) | 0.20 |
| Steroids + IVIG | 44 (38-50) | 61 (45-78) | 0.44 |
| IL-6 inhibitors (Tocilizumab or siltuximab) | 27 (8-43) | 4 (1-8) | **0.02** |
| IL-1Ra inhibitor (Anakinra) | 13 (10-17) | 5 (4-11) | **0.004** |
| **Outcome** |  |  |  |
| Deaths at the time of reporting | 2 (1-4) | 1 (0-3) | 0.21 |
| Duration of stay in days | 7.1 (5-7.9) | 9 (4.7-12.4) | 0.27 |

*Values stated as pooled estimate % (95% CI), unless specified
